# Saliva as a potential clinical specimen for diagnosis of SARS-CoV-2

**DOI:** 10.1101/2020.09.11.20192591

**Authors:** Debdutta Bhattacharya, Debaprasad Parai, Usha Kiran Rout, Rashmi Ranjan Nanda, Srikanta Kanungo, Girish Chandra Dash, Subrat Kumar Palo, Sidharth Giri, Hari Ram Choudhary, Jaya Singh Kshatri, Jyotirmayee Turuk, Bijay Kumar Mishra, Saroj Dash, Sanghamitra Pati

## Abstract

**Background:** It is almost nine months, still there is no sign to stop the spreading of the COVID-19 pandemic. Rapid and early detection of the virus is the master key to cease the rapid spread and break the human transmission chain. There are very few studies in search of an alternate and convenient diagnostic tool which can substitute nasopharyngeal swab (NPS) specimen for detection of SARS-CoV-2. We aimed to analyse the comparison and agreement between the feasibility of using the saliva in comparison to NPS for diagnosis of SARS-CoV-2.

**Methods:** A total number of 74 patients were enrolled for this study. We analysed and compared the NPS and saliva specimen collected within 48 h after the symptom onset. We used real time quantitative polymerase chain reaction (RT-qPCR), gene sequencing for the detection and determination SARS-CoV-2 specific genes. Phylogenetic tree was constructed to establish the isolation of viral RNA from saliva. We use Bland-Altman model to identify the agreement between two specimens.

**Findings:** This study shows a lower C_T_ mean value for the detection of SARS-CoV-2 ORF1 gene (27.07; 95% CI, 25.62 to 28.52) in saliva methods than that of NPS (28.24; 95% CI, 26.62 to 29.85) sampling method. Bland-Altman analysis produces relatively smaller bias and high agreement between these specimen tools. Phylogenetic analysis with the RdRp and Spike gene confirmed the presence of SARS-CoV-2 in the saliva samples.

**Interpretation:** In conclusion, our study highlights that saliva represents a promising tool in COVID-19 diagnosis and would reduce the exposure risk of frontline health workers which is one of biggest concern in primary healthcare settings.

## Introduction

Coronavirus disease 2019 (COVID□19) is an emerging infectious disease caused by severe acute respiratory syndrome coronavirus 2 (SARS-CoV-2) which was originated from Wuhan city, China in December 2019. Since then, COVID-19 became pandemic defining the current global health crisis and the greatest challenge that the world has faced since World War II. As on 2^nd^ September 2020, a total number of 25,602,665 confirmed cases has been registered and global death count reaches to 852,758. India is also fighting hard against the pandemic having a death report of 66,333 till date and still counting. Early detection, rapid test, contact trace following immediate isolation and treatment could only help us to restrict the COVID-19.^1-3^

Based on the current situation, it is urgently necessary to test more and more to contain the rapid spread of COVID-19 in the highly populated countries. Currently, sampling from nasopharyngeal swab (NPS) followed by RNA extraction and quantitative reverse-transcriptase polymerase chain reaction (RT-qPCR) is being used as the gold standard for the detection of SARS-CoV-2 infection.^4^ The conventional NPS testing always demands highly expertise personnel during sample collection unless it often leads to false negative results. Parallelly NPS collection technique is quite costly and time-consuming and often gives some uncomfortable complications like sneeze, cough or vomit to the sensitive patients.^5-6^ If we consider these factors, NPS is quite complicated to be robustly used in a highly populated and economically developing countries. Throat swab sample are also being used for the detection of COVID-19 but sometimes it’s treated as less sensitive method compared to NPS.^7-8^ Moreover, these two commonly used methods are invasive and very risky for the healthcare workers who are recruited and exposed during the collection of the samples.

Saliva is being secreted 90% from the major and rest from the minor salivary glands having pH around 6–7. It mainly consists with 99% water and remaining 1% contains organic molecules and inorganic salts.^9^ Saliva has been already proposed as an alternative diagnostic tool for the detection of other respiratory virus infection like influenza A and B, respiratory syncytial virus (RSV), SARS-CoV, coronaviruses HKU1, NL63, 229E and OC43.^10-12^ Recent studies have also shown that SARS-CoV-2 can be diagnosed from saliva of an asymptomatic persons and could also be transmitted by saliva droplets.^13^ Keeping all these in our mind, we have tried to test the feasibility of saliva as a non-invasive and safe alternate source of COVID-19 specimen. The study was carried out to assess the feasibility, acceptability of prospectively collecting saliva for detection of the presence of COVID-19 in saliva, and compare the pairing and agreement of the threshold cycle (C_T_) number with NPS specimen collected from the patients diagnosed with COVID-19.

## Materials and Methods

### Sample collection

A total of 74 suspected COVID-19 patients having mild to moderate symptoms were included in this study from different COVID care hospitals in Bhubaneswar, India. The study was approved by Institutional Ethics Committee and written consent was obtained from the patients. Both the NPS and saliva were collected within 48 h after symptoms onset. Saliva specimens were collected by the patients themselves in sterile sputum container and transported as per the standard guidelines by the Indian Council of Medical Research, New Delhi.

### Detection of SARS-CoV-2

The samples were subjected to detection of SARS-CoV-2 by Cobas 6800 instrument (Roche Molecular Diagnostics, Pleasanton, CA, USA). Two-target RT-PCR is used for the detection of SARS-CoV-2. The first target is an open reading frame 1 (ORF1) which is a non-structural region unique to SARS-CoV-2 and the second target is envelope E gene which is a structural protein conserved for the detection of pan-sarbecovirus. The tested sample was considered SARS-CoV-2 positive if both ORF1 and E genes or if only the ORF1 gene was found positive. Cycle threshold (C_T_) numbers were calculated by the inbuilt software of the system.

### RNA extraction, cDNA synthesis and gene sequencing

Total RNA was isolated from the SARS-CoV-2-infected saliva samples using QIAamp Viral RNA Mini Kit (Qiagen). The isolated RNA was reverse transcribed to cDNA using SuperScript IV First-Strand Synthesis System (Thermo Fisher Scientific, Carlsbad, USA) following the manufacturer’s protocol.^14^ In brief, 10 μL reaction mixture contained 5·5 μL of RNA, 0·5 μL of 10 mM deoxynucleotide triphosphate (dNTP) mixture, 2 μL SuperScript IV buffer (5X), 0·5 μL dithiothreitol (100 mM), 0·5 μL SuperScript IV reverse transcriptase (200U/μL), 0·5 μL of 50 μM random hexamers, DEPC-treated water to makeup volume. The mixture was initially incubated at 23°C for 10 min, then at 50°C for 10 min followed by 80°C for 10 min. Two genes *viz*. RNA-dependent RNA polymerase (*RdRp*) gene which is universal for all SARS-related coronaviruses and surface glycoprotein (*S*) gene of the SARS-CoV-2 were targeted for PCR and subsequent gene sequencing. Forward primer (5 - CAAGTGGGGTAAGGCTAGACTTT-3) and reverse primer (5 - ACTTAGGATAATCCCAACCCAT-3) were used to amplify a 344 bp sequence of RdRp gene. For the amplification of 156 bp long S gene, 5 - CCTACTAAATTAAATGATCTCTGCTTTACT-3 as forward and 5 - CAAGCTATAACGCAGCCTGTA-3 as reverse primers were chosen.^15^ Genes were amplified in Thermal Cycler Proflex (Applied Biosystems, USA) at 95°C for 10 min, followed by 50 cycles of 94°C for 1 min, 56°C for 1 min, and 72°C for 1 min. PCR products were run on the agarose gel and then the products were extracted by commercially available PureLink Quick Gel Extraction Kit (Invitrogen, USA) following manufacturer protocols. Purified PCR products were then sequenced with an ABI 3500xl Dx Genetic Analyzer (Applied Biosystems) following Sanger sequencing method using PCR primers. Two of the samples positive for the genes were subjected to sequencing & phylogenetic analysis to confirm the presence of SARS-CoV-2.

### Gene sequence and phylogenetic tree analysis

The forward and reverse sequences of RdRp and S genes isolated from saliva were edited manually in the electro-pherograms by SeqScape v2.5 software (Applied Biosystem, USA). Phylogenetic analysis was done using MEGA5.^16^ Kimura two parameter algorithm was used to calculate genetic distances and phylogenetic trees were constructed by the neighbour-joining (NJ) method. Bootstrap resampling method was used 1000 times to nullify any statistical errors and to reconstruct phylogenetic tree.

### Statistical analysis

Descriptive statistical analyses were performed using GraphPad Prism 7.04 for Windows (GraphPad Software, La Jolla, USA). Wilcoxon signed-rank test was used to compare NPS and saliva C_T_ values. Bland-Altman analysis was done to quantify the agreement between NPS and saliva by studying the mean C_T_ difference and constructing 95% limits of agreement. *p* values < 0.05 was considered as significant.

## Results

A total number of 53 subjects were tested positive both in NPS and saliva assay, whereas 5 samples were found only as NPS positive (**Table 1**). So, the specificity and sensitivity of this saliva based detection was determined as 100% and 91.37% respectively compared to samples collected as NPS. Only the positive samples (either from NPS or saliva) were compared for their respective C_T_ values. For ORF1 gene, 27 saliva samples showed either similar (C_T_±1.0) or lower C_T_ values compared to NPS samples whereas 31 saliva samples were found the same for E gene amplification. Among the positive samples, C_T_ values were similar between the two methods (*p* > 0.05) for detection of both the target genes. In NPS, the values ranged from 16.56 to 36.2 for ORF1 and 16.57 to 40.13 for E gene. The same was found to be from 16.83 to 34.65 (for ORF1) and 17.18 to 38.72 (for E gene) in case of saliva.

**Table 1:** Comparison of COVID-19 detection in RT-qPCR between NPS and saliva samples

| Nasopharyngeal swab |  | Positive | Negative |
| --- | --- | --- | --- |
| Saliva | Positive | 53 | 0 |
|  | Negative | 5 | 16 |

The pair wise comparison and phylogenetic tree analysis, bootstrap resampling and reconstruction revealed that both the branched with the SARS-CoV-2 isolated from different parts of the world **(Figure 1)**. The partial sequence of Sal1_SARS-CoV2_RMRCBB (GenBank Accession Number MT796264 and MT798848) and Sal2_SARSCoV2_RMRCBB (GenBank Accession Number MT796265 and MT798849) have been deposited into GenBank.

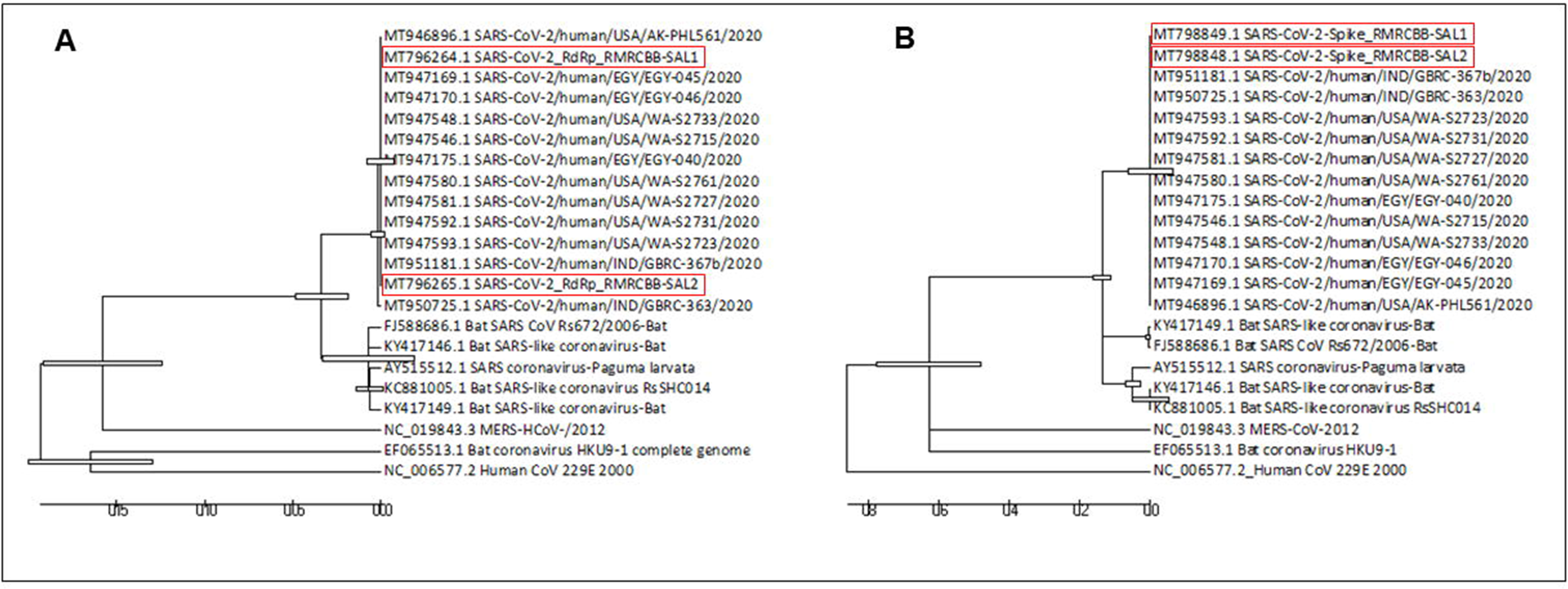

Statistically, we detected lower C_T_ value for ORF1 gene in the saliva specimens (mean C_T_ = 27.07; 95% confidence interval [CI], 25.62 to 28.52) than in the NPS specimens (mean C_T_ = 28.24; 95% CI, 26.62 to 29.85) (**Figure 2A**). The mean C_T_ value of E gene in saliva was found at 29.12 (at 95% CI, 27.46 to 30.79) which is almost similar to NPS samples (mean C_T_ = 29.04; 95% CI, 27.27 to 30.82) (**Figure 2B**).

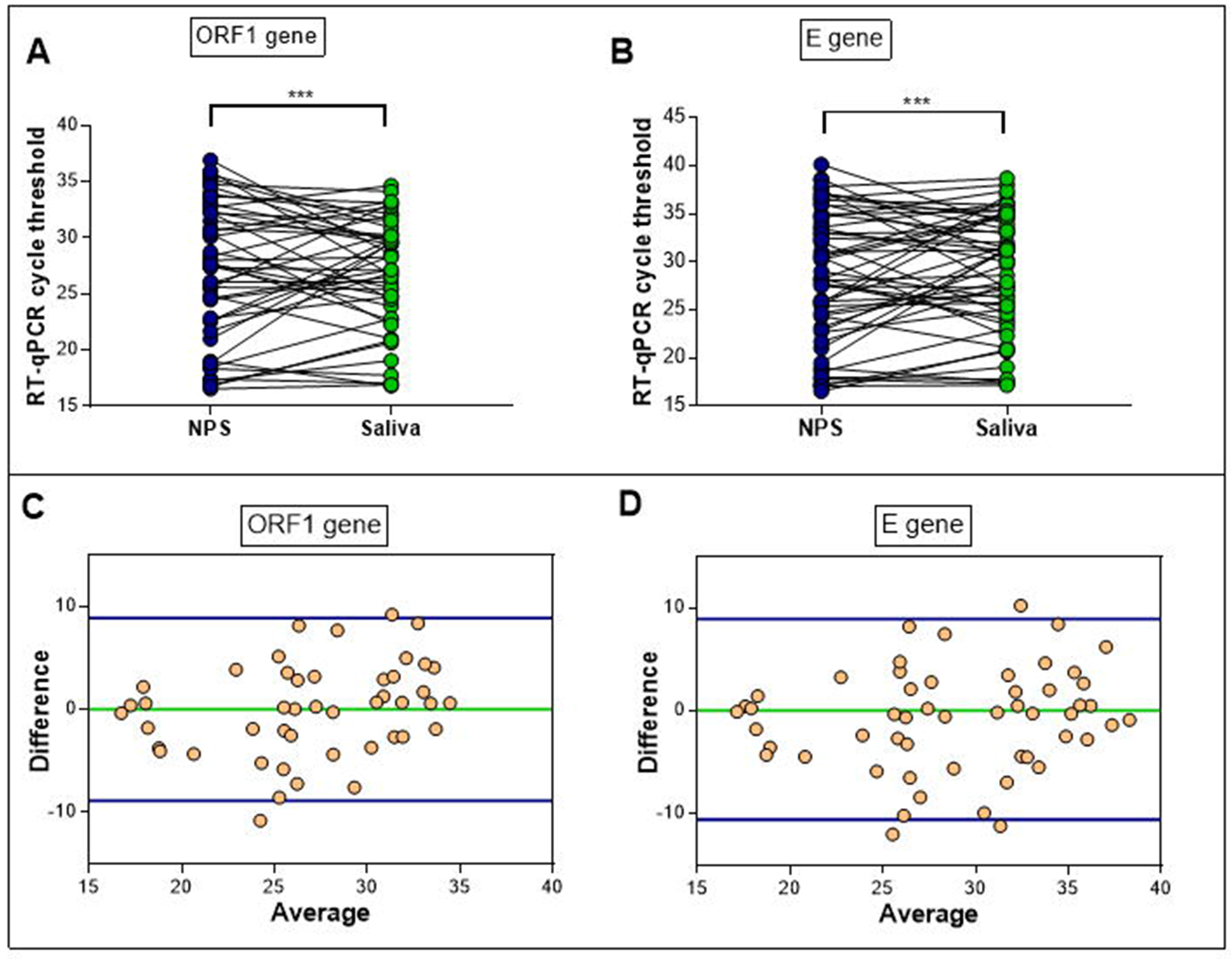

The difference of the two paired measurements is plotted against the mean of the NPS and saliva C_T_ values in Bland-Altman analysis. Bland-Altman analysis revealed a bias of 0.022 ± 4.53 (95% limits of agreement of −8.9 and 8.85) for ORF1, signifying close agreement (**Figure 2C**). A bias of 0.83 ± 4.98 (95% limits of agreement of −10.6 and 8.94) was determined for E gene when NPS and saliva methods were compared (**Figure 2D**).

## Discussion

Currently, NPS sample followed by RT-qPCR is considered as the gold standard test to diagnose SARS-CoV-2 from respiratory specimens.^4,17^ However, NPS isn’t an ideal and economical method for mass screening. It is quite time taking method which results into the crowding at specimen collection centres and subsequently putting the healthcare workers at high risk.^18^ Moreover, patients never find them in comfort zone during the NPS sample collection.^5,6^

The current study has shown that saliva can be a good alternative diagnostic tool for the detection of SARS-CoV-2. Other possible sources like sputum and oropharyngeal secretions are recently suggested for the molecular diagnosis of COVID19.^19^ Salivary droplets are already identified as the main source SARS-CoV-2 infection through human-to-human transmission below social distance limit of 2 meter.^20^ In our study, we tried to establish the feasibility of use of saliva as a potential specimen for diagnosis of SARS-CoV-2 as saliva can easily be collected by the patient and does not demand any trained personnel to collect it. The diagnosis of SARS-CoV-2 from saliva showed good sensitivity and specificity towards the detection of targeted genes by high throughput machine based on RT-qPCR. Statistical analyses found either similar (for E gene) or lower (for ORF1 gene) mean cycle threshold in saliva compared to NPS. Wilcoxon signed ranked test shows a highly significant pairing (*p*< 0.001) between the two sampling methods. As ORF1 is the confirmatory target for SARSCoV-2, it can be said that viral load in saliva is higher than NPS after symptoms onset. Gene sequencing analysis and subsequent phylogenetic tree construction also corroborated our automated RT-qPCR data and is the first report of SARS-CoV-2 isolates from saliva in India to the best of our knowledge. Overall, the NPS and saliva specimens show a high level of agreement among patients with a positive outcome in RT-qPCR. This allows the laboratories to collect COVID-19 testing sample not only from nasopharyngeal but also from saliva as a handy alternative. Our findings corroborate recently published results which show the presence of SARS-CoV-2 in saliva, however detailed and rigorous studies were not made available till now.^21-24^

Altogether, our study shows the reliability of saliva for the detection of COVID-19. Saliva samples can be collected by patient themselves leaving no complications as the process is fully non-invasive. This alternate could be a great relief for all the frontline health worker who all are engaged in the collection of NPS sample keeping themselves in a high risk. It could also be a worthwhile opportunity for the Government to reduce economic burden in this global crisis.

## Data Availability

The datasets used and/or analyzed during the current study are available from the corresponding author on reasonable request.

## Ethics approval and consent to participate

The study was cleared by institutional ethical committee.

## Author’s contribution

DB & SP designed the study. DB, DP, UKR, RRN, SD were involved in testing and data analysis. SK, GCD, SKP, HRC, JSK, JT and BM were responsible for data analysis and valuable inputs. DB, DP and SG did the statistical analysis. SP, DB, and DP wrote the manuscript. All authors have read and approved the final manuscript.

## Acknowledgement

The authors gratefully acknowledge all the healthcare workers for their tireless dedication at each level to fight COVID-19. The authors are thankful to ICMR for providing financial grants for this study.

## Reference

1 Pulla P. ‘The epidemic is growing very rapidly’: Indian government adviser fears coronavirus crisis will worsen. Nature 2020; 583: 180.

2 World Health Organization. 2020. Clinical management of Covid-19. Interim guidance. HO/2019-nCoV/clinical/2020.5

3 World Health Organization. 2020. Coronavirus Disease (COVID-19) dashboard. https://covid19.who.int/

4 Lippi G, Simundic AM, Plebani M. Potential preanalytical and analytical vulnerabilities in the laboratory diagnosis of coronavirus disease 2019 (COVID-19). Clin Chem Lab Med 2020; 58: 1070–6.

5 Marty FM, Chen K, Verrill KA. How to obtain a nasopharyngeal swab specimen. N Engl J Med 2020; 382: e76.

6 Qian Y, Zeng T, Wang H, et al. Safety management of nasopharyngeal specimen collection from suspected cases of coronavirus disease 2019. Int J Nurs Sci. 2020; 7(2): 153–156.

7 Wang H, Liu Q, Hu J, et al. Nasopharyngeal swabs are more sensitive than oropharyngeal swabs for COVID-19 diagnosis and monitoring the SARS-CoV-2 load. Front Med (Lausanne) 2020; 7: 334.

8 Zou L, Ruan F, Huang M, et al. SARS-CoV-2 viral load in upper respiratory specimens of infected patients. N Engl J Med 2020; 382: 1177–79.

9 Baghizadeh Fini M. Oral saliva and COVID-19. Oral Oncol 2020; 108: 104821.

10 Kim Y-g, Yun S G, Kim M Y, et al. Comparison between saliva and nasopharyngeal swab specimens for detection of respiratory viruses by multiplex reverse transcription-PCR. J Clin Microb 2017; 55: 226–33.

11 To KK, Lu L, Yip CC, et al. Additional molecular testing of saliva specimens improves the detection of respiratory viruses. Emerg Microbes Infect. 2017; 6: e49.

12 To KKW, Yip CCY, Lai CYW, et al. Saliva as a diagnostic specimen for testing respiratory virus by a point-of-care molecular assay: a diagnostic validity study. Clin Microbiol Infect 2019; 25: 372–78.

13 Xu R, Cui B, Duan X, et al. Saliva: potential diagnostic value and transmission of 2019-nCoV. Int J Oral Sci 12; 11.

14 Peiris JS, Lai ST, Poon LL, et al. Coronavirus as a possible cause of severe acute respiratory syndrome. Lancet 2003; 361: 1319–25.

15 Chan J F W, Yuan S, Kok K-H, et al. A familial cluster of pneumonia associated with the 2019 novel coronavirus indicating person-to-person transmission: a study of a family cluster. 2020. Lancet 2020; 395: 514–23.

16 Tamura K, Peterson D, Peterson N, et al. MEGA5: Molecular evolutionary genetics analysis using maximum likelihood, evolutionary distance, and maximum parsimony methods. Mol Biol Evol 2011; 28: 2731–39.

17 Wang Y, Kang H, Liu X, et al. Combination of RT-qPCR testing and clinical features for diagnosis of COVID-19 facilitates management of SARS-CoV-2 outbreak. J Med Virol 2020.

18 Nguyen LH, Drew DA, Graham MS et al. Risk of COVID-19 among front-line health-care workers and the general community: a prospective cohort study. Lancet Public Health 2020; 5: e475–83.

19 To KK, Tsang OT, Chik-Yan Yip C, et al. Consistent detection of 2019 novel coronavirus in saliva. Clin Infect Dis 2020 [Epub ahead of print].

20 Tian HY. 2019-nCoV: new challenges from coronavirua. Zhonghua Yu Fang Yi Xue Za Zhi 2020; 54: 235–8.

21 Azzi L, Carcano G, Gianfagna F, et al. Saliva is a reliable tool to detect SARS-CoV-2. J Infect 2020; 81: e45–e50.

22 Chen L, Zhao J, Peng J, et al. Detection of 2019-nCoV in saliva and characterization of oral symptoms in COVID-19 patients. Available at SSRN 3557140. 2020. https://ssrn.com/abstract=3556665

23 Pasomsub E, Watcharananan SP, Boonyawat K, et al. Saliva sample as a non-invasive specimen for the diagnosis of coronavirus disease 2019: a cross-sectional study. Clin Microbiol Infect. 2020; S1198-743X(20)30278-0.

24 Williams E, Bond K, Zhang B, et al. Saliva as a non-invasive specimen for detection of SARS-CoV-2. J Clin Microbiol. 2020: 58: e00776–20.

